# The effects of psilocybin therapy versus escitalopram on cognitive bias: A secondary analysis of a randomized controlled trial

**DOI:** 10.1101/2025.03.17.25324123

**Authors:** Jessica Henry, Bruna Giribaldi, David J Nutt, David Erritzoe, Robin Carhart-Harris, Taylor Lyons

**Affiliations:** Centre for Psychedelic Research, Imperial College London. Hammersmith Hospital, Du Cane Road, London, United Kingdom, W12 0NN; University of California, San Francisco. Parnassus Avenue, San Franscisco, CA 94143, United States

## Abstract

**Background:** Patients with Major Depressive Disorder (MDD) have more dysfunctional attitudes and pessimism than healthy individuals and these biases are correlated with depression severity. Psilocybin has demonstrated sustained remission in MDD.

**Methods:** Secondary analysis of a two-arm, randomized controlled trial (ClinicalTrials.gov Identifier: NCT03429075) assessing the effect of psilocybin therapy versus escitalopram on ‘maladaptive’ cognitive biases relevant to the construct of depression. Psilocybin group participants received two 25mg doses and escitalopram group received three weeks of daily 10mg, increased to 20mg for a following three weeks. Primary outcomes in this analysis were post-treatment changes in biases at six weeks compared with baseline, as measured using three validated psychological scales.

**Findings:** Fifty-nine MDD patients were randomly allocated to the psilocybin (n=30) or escitalopram (n=29) groups. Self-reported optimism showed a large and significant increase six-weeks after psilocybin treatment (*M*_*diff*_=6·63 *p*<0·0001; 95% *CI* [4·06, 9·20], *d=*1·1), whereas there was no change following escitalopram (*M*_*diff*_=1·52, *p*=0·205; 95% *CI* [−0·59, 3·62], *d=*0·4). Behavioral results found that patients were more optimistic about desirable life events after psilocybin treatment (*M*_*diff*_=0·16, *p*=0·0002; 95% *CI* [0·08, 0·23], *d=*1·1), but they were also less pessimistic about negative life events after escitalopram treatment (*M*_*diff*_=0·07, *p*=0·018; 95% *CI* [0·01, 0·13], *d=*0·5). We found improvements in all three domains of dysfunctional attitudes following psilocybin treatment: achievement (*M*_*diff*_=10·37, *p*<0·0001; 95% *CI* [6·38, 14·53], *d=*1·0); dependency (*M*_*diff*_=7·97, *p*<0·0001; 95% *CI* [4·00, 11·93], *d=*0·9) and self-control (*M*_*diff*_=6·40, *p*=0·0006; 95% *CI* [2·60, 10·20], *d=*0·8)), whereas only the achievement domain improved after escitalopram (*M*_*diff*_=4·10, *p*=0·005; 95% *CI* [1·35, 6·86], *d=*0·6).

**Interpretation:** These results suggest that two high-dose sessions with psilocybin therapy are superior to a six-week daily course of a selective serotonin-reuptake inhibitor antidepressant, in remediating negative cognitive biases in depression.

**Funding:** Supported by a private donation from the Alexander Mosley Charitable Trust and by the founding partners of Imperial College London’s Centre for Psychedelic Research.

**Author Contribution:** RCH conceptualized the study design and RCH and DJN oversaw the trial. TL plotted and analysed the data. TL and JH interpreted the data. JH wrote the manuscript and TL and RCH provided edits.

**Transparency Declaration:** The lead author and manuscript guarantor affirms that the manuscript is an honest, accurate, and transparent account of the study being reported; that no important aspects of the study have been omitted; and that any discrepancies from the study as planned (and, if relevant, registered) have been explained.

**Research Material Availability:** Research materials are available from the corresponding author, [JH], upon reasonable request.

## INTRODUCTION

Major Depressive Disorder (MDD) is among the leading causes of global disease burden and is associated with the highest number of days absent from work of any physical or mental disorder (1). Cognitive behavioural therapy (CBT) paired with selective serotonin reuptake inhibitors (SSRI) is the current recommended first-line treatment for MDD (2). Negative biases are argued to be a perpetuating cause of depressive symptoms and correction of such biases is central to CBT (3, 4). In support of the cognitive-bias model of MDD, extensive evidence shows that depressed patients have more dysfunctional attitudes and pessimism than healthy individuals, and that these biases are correlated with depression severity (5-7).

When twinned with psychological support, psilocybin has demonstrated rapid and sustained improvements in MDD in recent clinical trials, with 60-80% of patients having clinically significant reduction in depression at six-twelve months after treatment (8, 9). Primary psychoactive effects are mediated through agonism of the serotonin 2A receptor (5-HT_2A_R), a signalling pathway implicated in depression, mood states, and cognitive styles (10, 11). It has been hypothesised that impaired signalling at the 5-HT_2A_R may cause a rigid and pessimistic mind-set (5, 12), and that increased 5-HT_2A_R signalling may serve to enhance neural and psychological plasticity (10, 13-15). As such, increased 5-HT_2A_R signalling in the psychedelic state, when twinned with psychological support, can be utilized to promote adaptive revisions to these ‘biases’ (7). Building on previous research – which found less pessimistic and more accurate future forecasting after psilocybin therapy for treatment resistant depression (7) - here we investigated the impact of (two) psilocybin (doses) versus a six-week course of a commonly prescribed antidepressant, escitalopram, on cognitive biases (i.e., pessimistic future forecasting and dysfunctional attitudes) in patients with MDD. Assessed at baseline and a key six-week endpoint, we hypothesised a greater shift away from pessimistic forecasting after two treatment sessions with psilocybin therapy versus a six-week course of daily escitalopram.

## METHODS

### Trial Approvals

This study (ClinicalTrials.gov Identifier: NCT03429075) was carried out in accordance with Good Clinical Practice guidelines and received all necessary ethical and regulatory approvals. The authors assert that all procedures contributing to this work comply with the ethical standards of the relevant national and institutional committees on human experimentation and with the Helsinki Declaration of 1975, as revised in 2008. All procedures involving human subjects/patients were approved by the Brent Research Ethics Committee, U.K. Medicines and Healthcare Products Regulatory Agency, Health Research Authority, Imperial College London Joint Research and Compliance Office, and General Data Protection Regulations Office (REC reference 17/LO/0389). The National Institute for Health Research/Wellcome Trust Imperial Clinical Research Facility (ICRF) provided site-specific approvals. All participants provided written informed consent.

### Study Design and Participants

This was a two-arm, randomized controlled study. A total of fifty-nine patients with MDD were randomly allocated to the psilocybin (n=30) or escitalopram (n=29) treatment groups, matched for age, gender and education. Dosing regimens are depicted in figure 1. For more information on trial design, please refer to (16).

**Figure 1.**
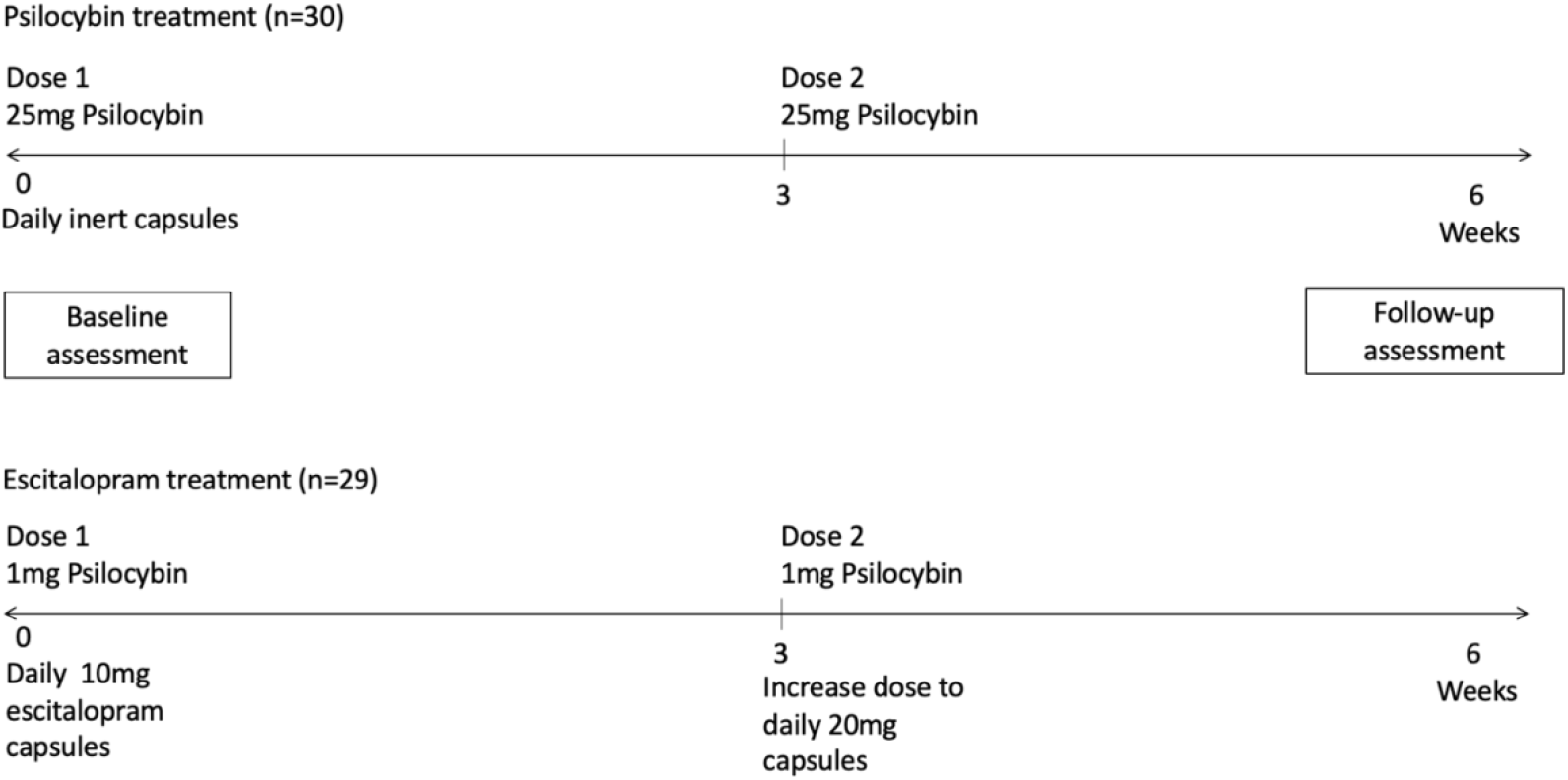
Schematic diagram of study design.

### Outcome Measures

The primary outcomes of the clinical trial have been discussed in (16). For the interest of this secondary analysis, the primary outcomes were post-treatment changes in cognitive biases (i.e., pessimism versus optimism bias and dysfunctional attitudes) at six weeks compared with baseline, as measured using three validated psychological scales.

We used the twenty four item *Dysfunctional Attitudes Scale* (DAS-24) to measure the presence and severity of dysfunctional beliefs on three common areas in depression: achievement, dependency and self-control (17). The *Revised Life Orientation Test* (LOT-R) (18) and *Prediction of Future Life Events* task (POFLE) (6) were used to assess changes in optimism versus pessimism biases. LOT-R is a self-reported ten item questionnaire developed to assess individual differences in generalised optimism versus pessimism (18). POFLE is a behavioural instrument used to objectively measure bias by assessing participant responses to forty potential future life events, twenty desirable (i.e. ‘positive’) and twenty undesirable (i.e. ‘negative’) events (see Strunk et al 2006 for scoring procedures) (6). POFLE is behavioural in the sense that it requires future forecasting rather than mere subjective ratings. POFLE was split into two versions, A and B, each containing twenty evenly balanced positive and negative items. All patients received both versions in balanced order; i.e., half of patients received POFLE version A at baseline and version B at follow-up, and the other half vice versa. Patients were contacted thirty days after completing each version to determine which of the events actually occurred.

To explore relationships between the primary outcomes and clinical improvements in mental health, we also assessed changes in depressive symptoms and psychological well-being at six weeks post-treatment from baseline. Depression severity was quantified using the *Beck Depression Inventory 1A* (BDI-1A)—a twenty one item questionnaire designed to assess characteristic attitudes and symptoms of depression (3). In this analysis, we chose to correlate measures of cognitive bias with BDI-1A, as the clinical trial analysis found a discrepancy between the Quick Inventory of Depression Symptomatology (QIDS) and other major efficacy outcome measures (16) (19). Well-being was measured using the *Flourishing Scale* (FS) (20) —an eight item questionnaire assessing domains of self-perceived success such as relationships, self-esteem and purpose. These scales were chosen due to the cognitive focus of the BDI-1A versus alternative depression rating scales, and the special relevance of the FS to the construct of optimism.

### Statistical Analysis

Statistical analyses were performed using SPSS version 27·0 (IBM Corp. Armonk, NY, USA) and illustrated using GraphPad Prism version 9·2 (GraphPad Software, La Jolla California USA). Data were subjected to t-tests, linear-mixed effects (LME) or two-way ANOVAs, as applicable. Welch’s correction was applied to comparisons with normative samples to control for unequal sample size and variance. Bonferroni-correction was applied to multiple *post hoc* comparisons. Relationships between the measures were assessed via correlations. Results provide 95% CIs around the mean differences and Cohen’s *d* effect sizes. Statistical significance was assumed at p≤0·050 probability level, at which the main clinical trial had 80% power to detect a difference between conditions in its primary outcomes.

## RESULTS

The number of the participants recruited and included in this analysis and the adverse events of the clinical trial have been discussed in (16).

### Cognitive Biases

#### Life Orientation Task (LOT-R)

A two-way mixed ANOVA examined the effect of time (i.e., baseline to six-weeks) and condition (i.e., psilocybin vs escitalopram) on cognitive bias, as measured by LOT-R (Figure 2A). A significant interaction was found between time and condition [*F*_(1,57)_=15·89, *p*=0·0002]. Subsequent *post-hoc* analyses of the within-subjects effects revealed a large (*d=*1·1) and significant increase in LOT-R scores following psilocybin (*M*_*diff*_=6.63, *SE*_*diff*_=1·26, *p*<0·0001; 95% *CI* [4·06, 9·20], *d=*1·1), but no difference following escitalopram (*M*_*diff*_=1·52, *SE*_*diff*_=0·70, *p*=0·205; 95% *CI* [−0·59, 3·62], *d=*0·4) at six-weeks relative to baseline. Analyses also found no between-subjects effects at baseline (*M*_*diff*_=0·55, *SE*_*diff*_=0·91, *p*=0·555; 95% *CI* [−2·39, 1·29], *d=*0·1), but a large (*d=*1·1) and significant difference at the six-week follow-up, with the psilocybin group having significantly greater LOT-R optimism scores than the escitalopram group (*M*_*diff*_=4·57, *SE*_*diff*_=1·17, *p*=0·0003; 95% *CI* [2·23, 6·91], *d=*1·0). The results suggest a superior treatment effect of psilocybin, with a significantly greater increase in LOT-R optimism scores observed following psilocybin (*M*=6·63, *SE*=1·09) compared with escitalopram (*M*=1·52, *SE*=0·65) treatment (*t*_(57)_=3·987, *p*=0·0002; 95% *CI* [2·55, 7·69], *d=*1·0).

**Figure 2.**
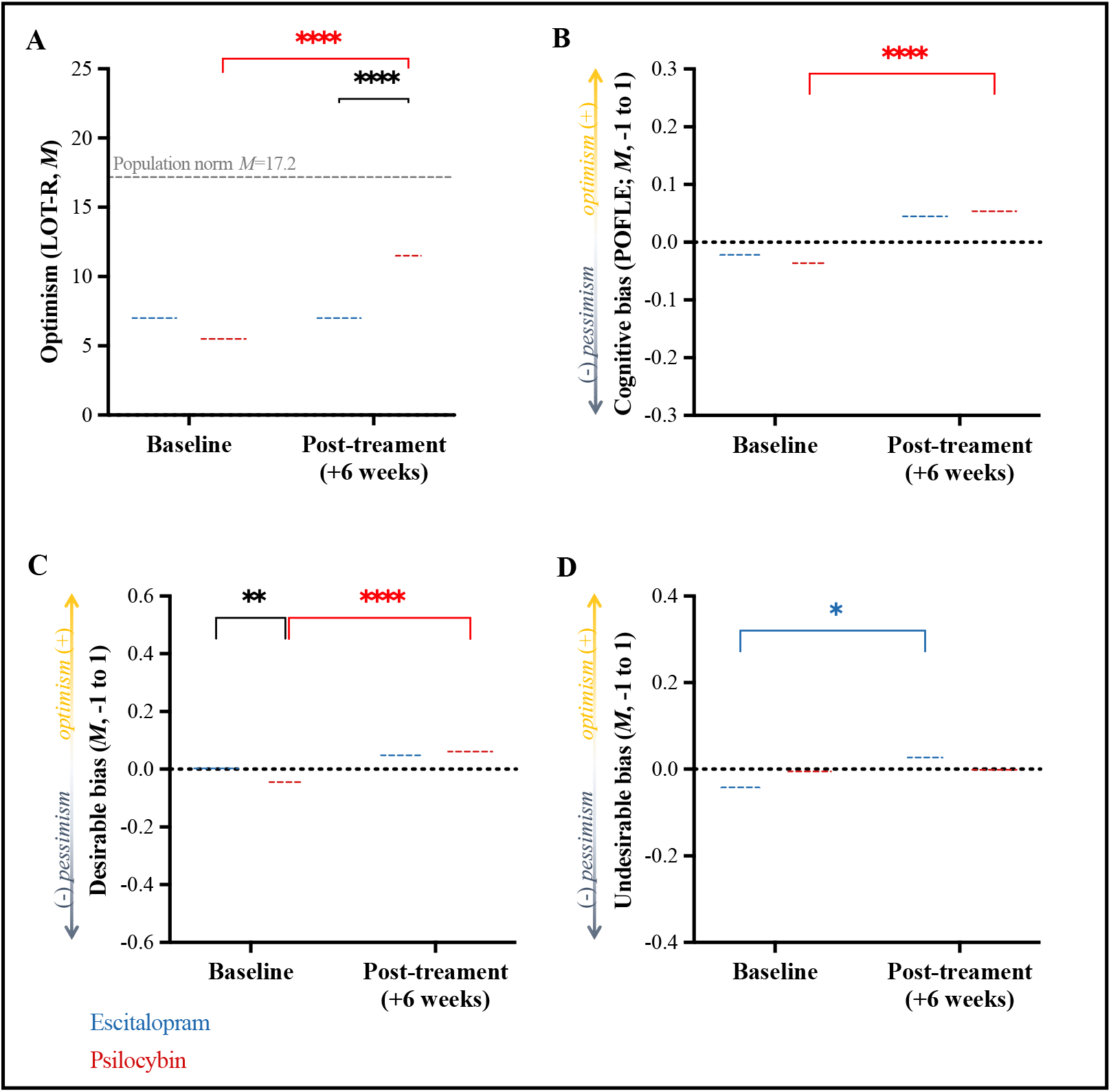
Cognitive Bias (A) LOT-R. Psilocybin group had significantly greater LOT-R scores 6 weeks after treatment (*M*_*diff*_=4.57, *SE*_*diff*_=1.17, *p*=0.0003; 95% CI [2.23, 6.91]) and significantly greater change scores at 6 weeks relative to baseline compared to the escitalopram group (*t*_(57)_=3.987, *p*=0.0002; 95% CI [2.55, 7.69]). **(B) POFLE**. A significant increase in bias scores was observed 6 weeks after psilocybin treatment relative to baseline (*M*_*diff*_=0.09, *SE*_*diff*_=0.02, *p*<0.0001; 95% CI [0.05, 0.14]). No within-group changes were observed following escitalopram treatment (*M*_*diff*_=0.04, *SE*_*diff*_=0.02, *p*=0.108; 95% CI [−0.01, 0.09]). **(C) POFLE: desirable events**. Psilocybin group displayed significantly greater pessimism than escitalopram group at baseline for desirable events (*M*_*diff*_=0.11, *SE*_*diff*_=0.04, *p*=0.007; 95% CI [0.03, 0.20]). Within-subjects analyses revealed a significant increase in bias scores for desirable events 6-weeks after psilocybin treatment compared to baseline (*M*_*diff*_=0.16, *SE*_*diff*_=0.04; *t*_(44)_=4.406, *p*=0.0002; 95% CI [0.08, 0.23]), with no significant changes following escitalopram treatment (*M*_*diff*_=0.04, *SE*_*diff*_=0.04; *t*_(18)_=0.989, *p*=0.336; 95% CI [−0.04, 0.11]). **(D) POFLE: undesirable events**. Escitalopram group showed a significant increase in undesirable bias scores at 6 weeks relative to baseline (*M*_*diff*_=0.07, *SE*_*diff*_=0.03, *p*=0.018; 95% CI [0.01, 0.13]). No significant changes were observed following psilocybin treatment (*M*_*diff*_=0.02, *SE*_*diff*_=0.02, *p*=0.672; 95% CI [−0.03, 0.08]). Data expressed as mean ± SE (*p* < 0.05*; *p* < 0.01**; *p* < 0.001***).

#### Prediction of Future Life Events (POFLE)

LME modelling examined the effect of time and condition on cognitive bias via the POFLE (Figure 2B). Analyses revealed a significant main effect of time (*F*_(1,99)_=20·86, *p*<0·0001), but no significant effect of condition (*F*_(1,99)_=0·86, *p*=0·355) or interaction (*F*_(1,99)_=2·78, *p*=0·098). *Post-hoc* comparisons showed no significant between-groups differences at baseline (*M*_*diff*_=0·04, *SE*_*diff*_=0·02, *p*=0·126; 95% *CI* [−0·09, 0·01], *d=*0·5) or six-week follow-up (*M*_*diff*_=0·01, *SE*_*diff*_=0·02, *p*>0·999; 95% *CI* [−0·04, 0·06], *d=*0·2). However, *post-hoc* comparisons of the within-subjects effects revealed a significant change in bias scores (i.e. decreased pessimism/increased optimism) at six-weeks relative to baseline following psilocybin (*M*_*diff*_=0·09, *SE*_*diff*_=0·02, *p*<0·0001; 95% *CI* [0·05, 0·14], *d=*0·8). No significant changes were observed following escitalopram (*M*_*diff*_=0·04, *SE*_*diff*_=0·02, *p*=0·108; 95% *CI* [− 0·01, 0·09], *d=*0·4).

As the POFLE contains both desirable (i.e., positive) and undesirable (i.e., negative) life events, subsequent analyses examined optimism vs. pessimism for events split by desirability (Figure 2C & 2D). Analyses revealed a significant time x condition interaction on forecasting of desirable events (*F*_(1,44)_=7·93, *p*=0·007). However, a significant between-groups difference in the forecasting of desirable events was found at baseline (*M*_*diff*_=0·11, *SE*_*diff*_=0·04, *p*=0·007; 95% *CI* [0·03, 0·20], *d=*0·8), whereby patients in the psilocybin group displayed significantly greater pessimism than the escitalopram group for desirable events.

To account for this baseline difference, the change in desirable bias from baseline was assessed using analysis of covariance (ANCOVA) with adjustment for baseline scores. After adjustment, there was no significant between-groups difference in the change in desirable event optimism/pessimism at six-weeks post-treatment (*F*_(1,44)_=0·50, *p*=0·482). This demonstrates that the baseline difference in forecasting of desirable events accounts for the between-groups variance in the change to follow-up and may occlude any actual between-condition differences in response on the POFLE. As such, subsequent analyses examined the within-condition effects. Analyses revealed a significant increase in optimism for desirable events six-weeks after psilocybin (*M*=0·07, *SD*=0·13) versus baseline (*M*=-0·09, *SD*=0·16; *M*_*diff*_=0·16, *SE*_*diff*_=0·04; *t*_(44)_=4·406, *p*=0·0002; 95% *CI* [0·08, 0·23], *d=*1·1), with no significant change at six-weeks following escitalopram treatment (*M*=0·04, *SD*=0·14) versus baseline (*M*=0·03, *SD*=0·12) (*M*_*diff*_=0·04, *SE*_*diff*_=0·04; *t*_(18)_=0·989, *p*=0·336; 95% *CI* [−0·04, 0·11], *d=*0·2).

No significant between-groups difference was observed for optimism/pessimism for undesirable events at baseline (Figure 2D). As such, LME analyses were performed. Analyses revealed a significant effect of time (*F*_(1,99)_=6·87, *p*=0·010), but no significant effect of condition (*F*_(1,99)_=0·47, *p*=0·496) or interaction (*F*_(1,99)_=1·76, *p*=0·187). *Post-hoc* comparisons showed no significant between-groups differences at baseline (*M*_*diff*_=0·04, *SE*_*diff*_=0·02, *p*=0·296; 95% *CI* [−0·02, 0·09], *d=*0·4) or six-week follow-up (*M*_*diff*_=0·01, *SE*_*diff*_=0·03, *p*>0·999; 95% *CI* [−0·07, 0·05], *d=*0·1). Analyses of the within-subjects effects found a significant improvement in pessimism scores for undesirable events (i.e. increased optimism) at six-weeks relative to baseline following escitalopram (*M*_*diff*_=0·07, *SE*_*diff*_=0·03, *p*=0·018; 95% *CI* [0·01, 0·13], *d=*0·5), but no significant changes following psilocybin (*M*_*diff*_=0·02, *SE*_*diff*_=0·02, *p*=0·672; 95% *CI* [−0·03, 0·08], *d=*0·2).

In summary, the results of the POFLE task suggest a differential treatment effect of psilocybin versus escitalopram. At the six-week post-treatment follow-up relative to baseline, patients in the psilocybin group displayed improvements in their optimism for positive/desirable events (*M*_*diff*_=0·16, *SE*_*diff*_=0·04; *t*_(44)_=4·406, *p*=0·0002; 95% *CI* [0·08, 0·23], *d=*0·1), with no change observed in their forecasting of negative/undesirable events (*M*_*diff*_=0·02, *SE*_*diff*_=0·02, *p*=0·672; 95% *CI* [−0·03, 0·08], *d=*0·2). On the contrary, the escitalopram group displayed improved pessimism for negative/undesirable events (*M*_*diff*_=0·07, *SE*_*diff*_=0·03, *p*=0·018; 95% *CI* [0·01, 0·13], *d=*0·5), with no change observed in their (generally pessimistic) forecasting of positive/desirable events (*M*_*diff*_=0·04, *SE*_*diff*_=0·04; *p*=0·336; 95% *CI* [−0·04, 0·11], *d=*0·3).

Contrary to the escitalopram-treated patients (*M*_*diff*_=0·04, *SE*_*diff*_=0·02, *p*=0·108; 95% *CI* [−0·01, 0·09], *d=*0·378), the psilocybin group also showed significantly greater optimism when forecasting future life events, overall (i.e. total scores not split by desirability); *M*_*diff*_=0·09, *SE*_*diff*_=0·02, *p*<0·0001; 95% *CI* [0·05, 0·14], *d=*0·8. This effect appeared to depend more on changes to their forecasting of desirable (*M*=0·16, *SE*=0·04) versus undesirable life events (*M*=0·06, *SE*=0·03). This overall post-treatment change in POFLE forecasting was greater after treatment with psilocybin than escitalopram (*t*_(44)_=2·049, *p*=0·046; 95% *CI* [0·002, 0·20], *d=*1·0).

#### Dysfunctional Attitudes Scale (DAS-24)

A two-way mixed ANOVA examined the effect of time and condition on dysfunctional attitudes, as measured by DAS-24 (Figure 3A). Analyses revealed significant time x condition interaction (*F*_(1,57)_=8·285, *p*=0·006). *Post-hoc* analyses found no difference between the groups at baseline (*M*_*diff*_=5·45, *SE*_*diff*_=5·38, *p*=0·316; 95% *CI* [5·33, 16·22], *d=*0·3), but a significant difference at follow-up—with the psilocybin group having significantly lower dysfunctional attitude scores relative to the escitalopram group (*M*_*diff*_=21·14, *SE*_*diff*_=5·63, *p*=0·0004; 95% *CI* [9·84, 32·42], *d=*1·0). Analyses of the within-subjects effects found a significant decrease in DAS-24 total scores in the psilocybin (*M*_*diff*_=-24·73, *SE*_*diff*_=4·46, *p*<0·0001; 95% *CI* [−33·86, − 15·61], *d=*1·0) and escitalopram (*M*_*diff*_=-9·03, *SE*_*diff*_=3·08, *p*=0·006; 95% *CI* [−15·34, −2·73], *d=*0·5) groups at six-weeks relative to baseline. To compare the degree of improvement over time, a between-groups t-test was conducted on the six-week DAS-24 total change scores from baseline. The results suggest a superior treatment effect of psilocybin, with a significantly greater improvement in DAS-24 scores following psilocybin (*M*=-24·73, *SE*=4·46) compared with escitalopram (*M*=-9·03, *SE*=3·08) treatment (*M*_*diff*_=-15.70, *SE*_*diff*_=5.45, *t*_(57)_=2·878, *p*=0·006; 95% *CI* [−26·62, −4·78], *d=*0·8).

**Figure 3.**
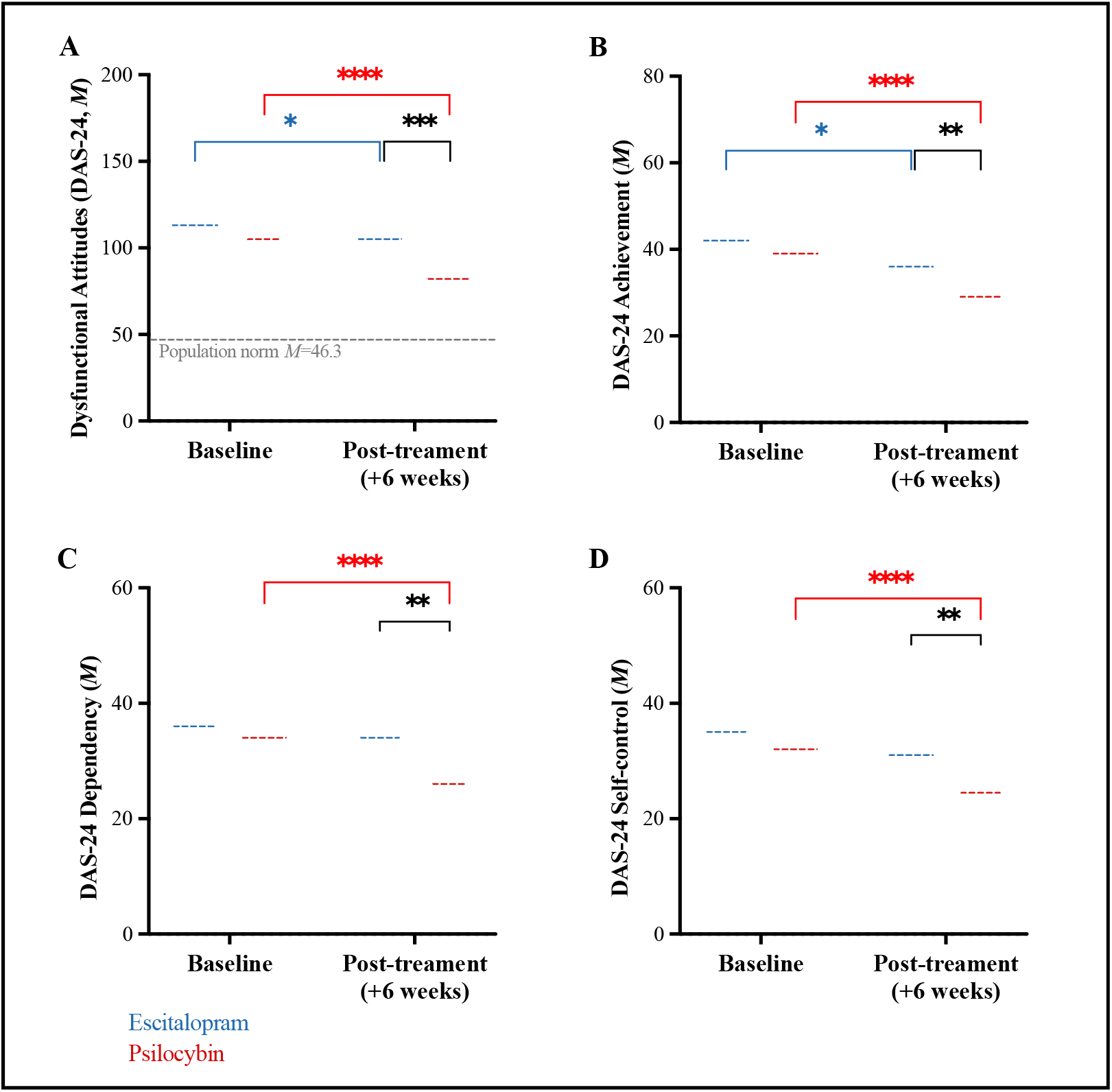
Dysfunctional Attitudes. **(A)** Total **DAS-24**. Psilocybin group had significantly lower scores at follow up compared to escitalopram group (*M*_*diff*_=21.14, *SE*_*diff*_=5.63, *p*=0.0004; 95% CI [9.84, 32.42]). Psilocybin group had significantly greater change at 6 weeks from baseline compared to escitalopram treatment group (*t*_(57)_=2.878, *p*=0.006; 95% CI [−26.62, −4.77]). **(B) Achievement**. Psilocybin scores were significantly lower than escitalopram scores at follow up (*M*_*diff*_=8.09, *SE*_*diff*_=2.50, *p*=0.002; 95% CI [3.09, 13.10]). **(C) Dependency**. Psilocybin scores were significantly lower than escitalopram scores at follow up (*M*_*diff*_=7.07, *SE*_*diff*_=1.96, *p*=0.001; 95% CI [3.13, 11.00]). **(D) Self-control**. Psilocybin scores were significantly lower than escitalopram scores at follow up (*M*_*diff*_=5.98, *SE*_*diff*_=1.93, *p*=0.003; 95% CI [2.11, 9.85]). Data expressed as mean ± SE (*p* < 0.05*; *p* < 0.01**; *p* < 0.001***).

Next, a two-way mixed ANOVA examined the effect of time and DAS-24 subscale scores (i.e. achievement, dependency and self-control; Figure 3B-D). Analyses revealed a significant time x subscale interaction (*F*_(2,144)_=4·827, *p*=0·010). *Post-hoc* analyses found no between-groups differences on the achievement (*M*_*diff*_=1·83, *SE*_*diff*_=2·43, *p*=0·456; 95% *CI* [−3·06, 6·72], *d=*0·2; Figure 3B), dependency (*M*_*diff*_=1·65, *SE*_*diff*_=1·73, *p*=0·345; 95% *CI* [−1·82, 5·13], *d=*0·2; Figure 3C) or self-control (*M*_*diff*_=1·96, *SE*_*diff*_=1·93, *p*=0·314; 95% *CI* [−1·90, 5·83], *d=*0·3; Figure 3D) scores at baseline. Yet at the six-week follow-up, a significant between-groups difference was found on all three subscales, with significantly lower scores following psilocybin compared with escitalopram on all subscales: achievement (*M*_*diff*_=8·09, *SE*_*diff*_=2·50, *p*=0·002; 95% *CI* [3·09, 13·10], *d=*0·8); dependency (*M*_*diff*_=7·07, *SE*_*diff*_=1·96, *p*=0·001; 95% *CI* [3·13, 11·00], *d=*0·9); self-control (*M*_*diff*_=5·98, *SE*_*diff*_=1·93, *p*=0·003; 95% *CI* [2·11, 9·85], *d=*0·8). Within-subjects analyses revealed a significant decrease in achievement scores (*M*_*diff*_=4·10, *SE*_*diff*_=1·34, *p*=0·005; 95% *CI* [1·35, 6·86], *d=*0·6) in the escitalopram group at six-weeks relative to baseline (Figure 3B), yet no significant changes were observed on dependency (*M*_*diff*_=2·55, *SE*_*diff*_=1·09, *p*=0·081; 95% *CI* [−0·23, 5·33], *d=*0·4; Figure 3C) or self-control (*M*_*diff*_=2·28, *SE*_*diff*_=1·36, *p*=0·271; 95% *CI* [−1·08, 5·83], *d=*0·3; Figure 3D). As implied by the between-condition difference, scores were significantly decreased on all three subscales in the psilocybin group at six-weeks relative to baseline: achievement (*M*_*diff*_=10·37, *SE*_*diff*_=1·95, *p*<0·0001; 95% *CI* [6·38, 14·53], *d=*1·0; Figure 3B); dependency (*M*_*diff*_=7·97, *SE*_*diff*_=1·56, *p*<0·0001; 95% *CI* [4·00, 11·93], *d=*0·9; Figure 3C) and self-control (*M*_*diff*_=6·40, *SE*_*diff*_=1·49, *p*=0·0006; 95% *CI* [2·60, 10·20], *d=*0·8; Figure 3D).

Thus, escitalopram treatment appeared to improve dysfunctional attitudes related to *achievement* but not *dependency* or *self-control*. In contrast, the results showed an improvement in all sub-areas of dysfunctional attitudes following psilocybin treatment.

### Depressive symptoms and well-being

Two-way mixed ANOVAs were conducted to examine the effect of time and condition on secondary outcomes.

#### Beck Depression Inventory 1A (BDI-1A)

Analyses of the BDI-1A revealed a significant time x condition interaction (*F*_(1,57)_=6·821, *p*=0·011). *Post-hoc* analyses found no difference between the groups at baseline (*M*_*diff*_=0·38, *SE*_*diff*_=2·40, *p*>0·999; 95% *CI* [−5·07, 5·83], *d*<0·1), but a significant difference at the six-week primary endpoint —with the psilocybin group having significantly lower BDI-1A scores relative to the escitalopram group (*M*_*diff*_=7·19, *SE*_*diff*_=2·40, *p*=0·007; 95% *CI* [−1·75, −12·64], *d=*0·7; Figure 4A). Within-subjects analyses found a significant decrease in BDI-1A scores following psilocybin (*M*_*diff*_=18·40, *SE*_*diff*_=2·24, *p*<0·0001; 95% *CI* [13·72, 23·08], *d=*1·5) and escitalopram (*M*_*diff*_=10·83, *SE*_*diff*_=1·83, *p*<0·0001; 95% *CI* [6·07, 15·59], *d=*1·1) at six-weeks relative to baseline. As implied by the interaction, a between-groups t-test on six-week BDI- 1A change scores from baseline to week-six showed a significantly greater decrease in depressive symptom severity following psilocybin (*M*=-18·40, *SE*=2·24) than escitalopram (*M*=-10·83, *SE*=1·83) treatment (*M*_*diff*_=-7.57, *SE*_*diff*_=2.899, *t*_(57)_=2·612, *p*=0·012; 95% *CI* [− 13·38, −1·77], *d=*0·7).

**Figure 4.**
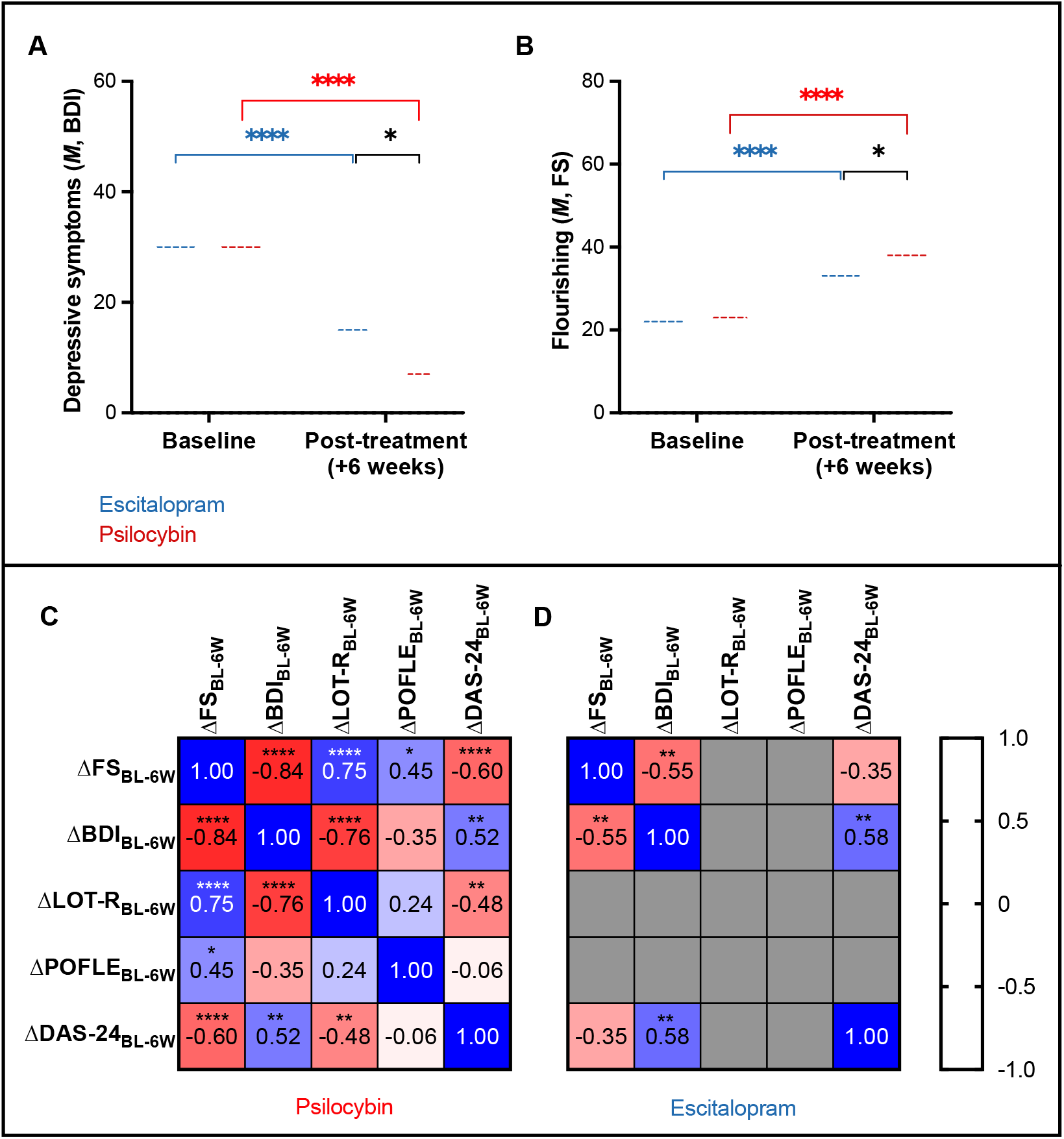
Secondary outcomes. (A) Depressive symptoms. Analyses revealed a significant decrease in BDI scores at 6 weeks post-treatment relative to baseline in both conditions (*psilocybin group*: *M*_*diff*_=18.40, *SE*_*diff*_=2.24, *p*<0.0001; 95% CI [13.72, 23.08]; *escitalopram group*: *M*_*diff*_=10.83, *SE*_*diff*_=1.83, *p*<0.0001; 95% CI [6.07, 15.59]). There was a significantly greater decrease in BDI scores following psilocybin treatment compared to escitalopram (*t*_(57)_=2.612, *p*=0.011; 95% CI [−13.38, −1.77], d=11.1). **(B) Psychological well-being**. Significantly increased FS scores were observed in both conditions at 6-weeks relative to baseline (*psilocybin group*: *M*_*diff*_=14.43, *SE*_*diff*_=1.83, *p*<0.0001; 95% CI [10.23, 18.64]; *escitalopram group*: *M*_*diff*_=8.93, *SE*_*diff*_=1.86, *p*<0.0001; 95% CI [4.65, 13.21]). A significantly greater improvement in FS scores was observed following psilocybin compared to escitalopram (*M*_*diff*_=5.57, *SE*_*diff*_=2.89; *t*_(57)_=2.110, *p*=0.039; 95% CI [−10.72, −0.28], d=10.0).. Data expressed as mean ± SE (*p* < 0.05*; *p* < 0.01**; *p* < 0.001***). **(C) Psilocybin correlations**. There was a significant correlation found between the change in BDI scores with DAS-24 (*r*_*s*_=0.519, *p*=0.003; 95% CI [0.18, 0.75]) and LOT-R (*r*_*s*_=-0.758, *p*<0.0001; 95% CI [−0.88, −0.54]) scores following psilocybin treatment. The negative correlation between the change in BDI and POFLE scores did not reach statistical significance (*r*_*s*_=-0.349, *p*=0.074; 95% CI [−0.65, 0.05]). A significant correlation was found between the change in FS and all primary outcomes following psilocybin treatment—i.e. LOT-R (*r*_*s*_=0.753, *p*<0.0001; 95% CI [0.53, 0.88]), POFLE (*r*_*s*_=0.448, *p*=0.019; 95% CI [0.07, 0.71]) and DAS-24 (*r*_*s*_=-0.599, *p*<0.0001; 95% CI [−0.79, −0.29]). **(D) Escitalopram correlations**. There was a significant negative correlation found between change in BDI and DAS-24 scores following escitalopram treatment (*r*_*s*_=0.579, *p*=0.001; 95% CI [0.26, 0.78]). There was no relationship found between the change in FS and DAS-24 scores following escitalopram treatment. No significant post-escitalopram changes were observed in cognitive bias (via the LOT-R and POFLE) for correlational analyses with BDI or FS.

#### Flourishing Scale (FS)

Analyses of psychological flourishing using the FS revealed a significant time x condition interaction (*F*_(1,57)_=4·454, *p*=0·039). *Post-hoc* analyses found no difference between the groups at baseline (*M*_*diff*_=0·16, *SE*_*diff*_=2·19, *p*>0·999; 95% *CI* [−4·84, 5·15], *d<*0·1), but a significant difference at the six-week endpoint —with significantly greater flourishing scores following psilocybin relative to escitalopram (*M*_*diff*_=5·35, *SE*_*diff*_=2·19, *p*=0·039; 95% *CI* [−10·34, −0·35], *d=*0·6; Figure 4B). Within-subjects analyses found significantly increased flourishing following psilocybin (*M*_*diff*_=14·43, *SE*_*diff*_=1·83, *p*<0·0001; 95% *CI* [10·23, 18·64], *d=*1·7) and escitalopram (*M*_*diff*_=8·93, *SE*_*diff*_=1·86, *p*<0·0001; 95% *CI* [4·65, 13·21], *d=*1·4) at six-weeks relative to baseline. In line with the interaction effect, a between-groups t-test revealed a significantly greater change (increase) in flourishing following psilocybin (*M*=14·43, *SE*=2·28) than escitalopram (*M*=8·93, *SE*=1·21) treatment (*M*_*diff*_=7·57, *SE*_*diff*_=2·89; *t*_(57)_=2·110, *p*=0·039; 95% *CI* [−10·72, −0·28], *d=*0·6).

### Relationships between primary and secondary outcomes

Next, we explored relationships between the change in primary and secondary outcomes at the six-week endpoint.

#### Relationship between cognitive biases and depression severity

First, we assessed the relationship between the change in cognitive bias and depression severity. A correlation was found between changes in DAS-24 and BDI-1A scores following psilocybin (*r*_*s*_=0·519, *p*=0·003; 95% CI [0·18, 0·75]; Figure 4C) and escitalopram (*r*_*s*_=0·579, *p*=0·001; 95% CI [0·26, 0·78]; Figure 4D), such that a greater decrease in dysfunctional attitudes correlated with a greater decrease in depressive symptoms in both conditions. Similarly, we found a negative correlation between changes LOT-R and BDI-1A scores following psilocybin (*r*_*s*_=-0·758, *p*<0·0001; 95% CI [−0·88, −0·54]), such that a greater increase in optimism correlated with a greater decrease in depressive symptoms (Figure 4C). To the contrary, the negative correlation between changes in POFLE and BDI-1A scores following psilocybin did not reach statistical significance (*r*_*s*_=-0·349, *p*=0·074; 95% CI [−0·65, 0·05]; Figure 4C). Consistent with the lack of significant change in pessimism-optimism bias following escitalopram treatment (i.e., LOT-R and total POFLE values) (see Figure 2A-B), relationships between these values and BDI-1A change scores were not significant.

#### Relationship between cognitive biases and well-being

Next, we assessed the relationship between the change in the main cognitive bias outcomes and ‘flourishing’. A correlation was found between the change in FS and all three cognitive bias outcomes following psilocybin—i.e. LOT-R (*r*_*s*_=0·753, *p*<0·0001; 95% CI [0·53, 0·88]), POFLE (*r*_*s*_=0·448, *p*=0·019; 95% CI [0·07, 0·71]) and DAS-24 (*r*_*s*_=-0·599, *p*<0·0001; 95% CI [−0·79, −0·29])—such that a greater improvement in flourishing correlated with greater improvements in both pessimism-optimism biases and dysfunctional attitudes (Figure 4C). To the contrary, no relationship was found between the change in DAS-24 (*r*_*s*_=-0·350, *p*=0·063; 95% *CI* [−0·64, 0·03]) and FS scores following escitalopram (Figure 4D)—and no significant post-escitalopram changes were observed in pessimism-optimism bias (via the LOT-R and total POFLE) for correlation analyses with FS (see Figure 2A-B).

## DISCUSSION

The present study assessed the effect of psilocybin versus escitalopram on various indices of ‘maladaptive’ cognitive biases relevant to the construct of depression. We found significantly increased self-reported optimism six-weeks after psilocybin treatment, with no change following escitalopram. Behavioral results largely supported these subjective data, but with some nuance– i.e., patients were more optimistic about the likelihood of experiencing positive life events after treatment with psilocybin, but they were also less pessimistic about the likelihood of experiencing negative life events after treatment with escitalopram. We found improvements in all sub-domains of dysfunctional attitudes following psilocybin treatment – but only one of them after escitalopram. These results suggest that two high-dose sessions with psilocybin therapy are superior to a six-week daily course of a selective serotonin-reuptake inhibitor antidepressant, in remediating negative cognitive biases in depression.

### Does psilocybin therapy ‘correct’ cognitive biases integral to depression?

To the best of our knowledge, this is the first study to directly address the impact of psilocybin therapy on behavioural indices of dysfunctional attitudes. The reduction of dysfunctional attitudes found here after escitalopram is consistent with previous reports on the effects of SSRIs on this construct (21). We found a greater magnitude of change for overall dysfunctional attitudes and across all sub-domains following psilocybin treatment. These findings indicate that psilocybin therapy positively impacts a broader range of attitude-related dysfunctionality than escitalopram.

We also found increased self-reported optimism six-weeks after psilocybin treatment, with no change following escitalopram. These findings are consistent with previous reports of sustained increases in optimism following psychedelic compounds (22). In a previous study of ours, psilocybin therapy was found to correct severe pessimism biases in patients with treatment-resistant depression and improve the accuracy of their future forecasting—i.e., post-treatment, future forecasting aligned better with their actual life experiences (7). Notably, in this previous work, psilocybin therapy was shown to specifically target pessimism bias for *desirable* life events (7), consistent with the current finding that psilocybin and escitalopram differentially treat expectations regarding desirable and undesirable life events, respectively. That is, here we found more optimistic expectations for desirable life events following psilocybin therapy, but reduced expectations for undesirable life events following escitalopram. Evidence suggests that SSRIs can flatten affective responsivity, a phenomenon known as ‘emotional blunting’ (23). We speculate that the decrease in pessimism for undesirable life events via escitalopram may relate to a muting of affective tone. Evidence of a putative valence specificity in the action of SSRIs, as seen here via escitalopram and the POFLE data, may in fact relate to a greater capacity for an effect in the negative domain. This reasoning would explain why escitalopram was unable to enhance forecasting of desirable life events (i.e., because it blunts affective tone and doesn’t enhance hedonic tone (24)) – however, it does not necessarily explain the lack of effect of psilocybin on forecasting negative life events.

Extensive evidence highlights the role of serotonergic mechanisms in mood states and cognitive styles (10). The 5-HT_2A_R – the key receptor through which psychedelics elicit their signature effects – is centrally involved in cognitive modulation, with implications for affective processing (11). Serotonin efflux and 5-HT2AR signalling specifically, have been associated with upregulated neuroplasticity (13), learning rate (14), cognitive flexibility (10) (15), psychological flexibility (25), and sensitivity to context (8, 25). 5-HT2ARs have been found to be upregulated in unmedicated MDD (12), high trait neuroticism (26) and, particularly, in depressed individuals with dysfunctional attitudes - such that apparent receptor concentrations correlate with the severity of dysfunction (5).

A predictive processing framework has been used to explain a plethora of states and traits in cognitive neuroscience and psychiatry – including the psychedelic state and action of psychedelic therapy (27). According to one model, psychedelics act acutely to decrease the ‘precision weighing’ of predictive models encoded in brain activity and circuitry (27) – an effect observable via increases in the entropy of (model-encoding) spontaneous brain activity (28), synonymous with a dysregulation of statistical regularities within the basal activity. Extending on this, a common feature of mental illness may be an excessive reinforcement of maladaptive circuitry – or an excessive ‘precision-weighting’ of pathological beliefs (27) – synonymous with excess regularities in the spontaneous activity, as laid out in detail here (4).

Thus, the ability of psychedelics to affect maladaptively reinforced patterns of thought and behaviour, by dysregulating the patterns of brain activity upon which they rest, may be a core aspect of their therapeutic potential (4). Accumulating evidence indicates that psychedelic therapy with classic 5-HT_2A_R agonists may be effective for treating anxiety, depression, obsessive-compulsive disorder, addiction, and distress caused by serious illness (8, 9, 16, 29) (30). However, all of these studies have involved positive contextual manipulations and are highly sensitive to patient and experimenter confirmation biases. Consistent with the so-called ‘plasticity paradox’ i.e., that enhanced plasticity is not inherently salutogenic or healing, future work is required to examine the question of whether psychedelics could also be used to enhance the learning of cognitive biases – if e.g., contextual conditions were manipulated in a way to promote such learning (27). The ethics and potential risks of psychedelic research, psychedelic therapy and general psychedelic-use are relevant in this regard.

### Limitations

Limitations of the present trial design have been discussed before (16) and thus, will only be briefly covered here. We did not assess blinding integrity. Moreover, due to the subjective drug effects of psilocybin and side effects of SSRIs, we suspect that most participants and the researchers could correctly guess treatment allocations. Another limitation of this study is that SSRIs take several weeks to reach full therapeutic effect and our study design of 6 weeks may not have allowed sufficient time for escitalopram to achieve its maximum benefit. Despite active efforts to recruit a diverse study population, our patients were majority Caucasian, highly educated males with baseline moderate depression scores. This limits the generalisability of results and further work must be done to improve diversity in recruiting patient populations for psychedelic research. Lastly, our present analyses corrected for multiple comparisons within measures (e.g., sub-factors) but not between them, and we recognize that several metrics have been employed and analyzed in this study. However, given the explorative nature of the trial, we feel that this approach can justified for inspiring 1) hypothesis generation - including nuanced perspectives on potential treatment mechanisms, as well as 2) focused consideration on the important construct of dysfunctional attitudes and cognitive biases in depression, which might otherwise be glossed over.

## Conclusions

Here we have assessed various measures of negative cognitive bias in patients with major depressive disorder treated with either psilocybin therapy or escitalopram. Psilocybin therapy appeared to have a more robust and comprehensive positive effect on these biases – inspiring inferences on its therapeutic mechanisms and lending further support to efforts to scrutinize its safety and efficacy at greater scale.

## Data Availability

The data that support the findings of this study are available from the corresponding author, [JH], upon reasonable request.

## Acknowledgements

We thank Ms Bruna Giribaldi for trial coordination and data collection; Dr. Rosalind Watts, Ms. Michelle Baker-Jones, Ms Ashleigh Murphy-Beiner, Dr. Roberta Murphy, Dr Jonny Martell, and Dr David Erritzoe for clinical oversight and guide roles; Ms Renee Harvey, Dr. Graham Campbell, Dr. Benjamin Waterhouse, Dr. Frederico Magalhães, Mr. James Close, Dr. Leor Roseman, Ms. Hilary Platt, Mr. Gregory Donaldson, and Dr. Chris Timmermann for their voluntary assistant guide roles; Dr. Meg Spriggs and Ms. Laura Kärtner for their support with data collection; Dr. Louise Paterson and Dr. Robin Tyacke for their assistance with randomisation, blinding, and drug accountability; Ms. Ghazel Mukhtar for her assistance with administrative tasks; and Dr. Tim Read for his clinical supervision and guidance.

## References

1. Kessler RC. The costs of depression. The Psychiatric Clinics of North America. 2012;35(1):1–14.

2. NICE. Depression in adults: recognition and mangement National Institute of Care and Excellence 2009.

3. Beck AT, Rush AJ, Shaw BF, Emery G. Cognitive therapy of depression. New York: Guildford Press. 1979.

4. Carhart-Harris RL, Chandaria S, Erritzoe DE, Gazzaley A, Girn M, Kettner H, et al. Canalization and plasticity in psychopathology. Neuropsychopharmacology. 2023;226(109398).

5. Meyer JH, McMain S, Kennedy SH, Korman L, Brown GM, DaSilva JN, et al. Dysfunctional attitudes and 5-HT2 receptors during depression and self-harm. The American Journal of Psychiatry. 2003;160(1):90–9.

6. Strunk DR, Lopez H, DeRubeis RJ. Depressive symptoms are associated with unrealistic negative predictions of future life events. Behaviour Research and Therapy. 2006;44(6) 861–82.

7. Lyons T, Carhart-Harris RL. More Realistic Forecasting of Future Life Events After Psilocybin for Treatment-Resistant Depression. Frontiers in Psychology. 2018;9(1721).

8. Carhart-Harris RL, Bolstridge M, Day CMJ, Rucker J, Watts R, Erritzoe DE, et al. Psilocybin with psychological support for treatment-resistant depression: six-month followup. Psychopharmacology. 2018;235(2):399–408.

9. Ross S, Bossis A, Guss J, Agin-Liebes G, Malone T, Cohen B, et al. Rapid and sustained symtpom reduction following psilocybin treatment for anxiety and depression in patients with life-threatening cancer: a randomized controlled trial. Journal of Psychopharmacology (Oxford, England). 2016;30(12) 1165–80.

10. Carhart-Harris RL, Nutt DJ. Serotonin and brain function: a tale of two receptors. Journal of Psychopharmacology (Oxford, England). 2017;31(9):1091–120.

11. Kometer M, Schmidt A, Bachmann R, Studerus E, Seifrtiz E, Vollenweider FX. Psilocybin biases facial recognition, goal-directed behaviour, and mood state toward positive relative to negative emotions through different serotonergic subreceptors Biological Psychiatry. 2012;72(11):898–906.

12. Bhagwagar Z, Hinz R, Taylor M, Fancy S, Cowen P, Grasby P. Increased 5-HT(2A) receptor binding in euthymic, medication-free patients recovered from depression: a positron emission study The American Journal of Psychiatry. 2006;163(9):1580–7.

13. Ly C, Greb AC, Cameron LP, Wong JM, Barragan EV, Wilson PC, et al. Psychedelics Promote Structural and Functional Neural Plasticity. Cell Reports. 2018;23(11):3170–82.

14. Kanen J, Luo Q, Rostami Kandroodi M, Cardinal R, Robbins T, Carhart-Harris RL, et al. Effect of lysergic acid diethylamide (LSD) on reinforcement learning in humans. 2020.

15. Matias S, Lottem E, Dugué GP, Mainen ZF. Activity patterns of serotonin neurons underlying cognitive flexibility. Elife. 2017;6(e20552).

16. Carhart-Harris RL, Giribaldi B, Watts R, Baker-Jones M, Murphy-Beiner A, Murphy R, et al. Trial of Psilocybin versus Escitalopram for Depression. The New England Journal of Medicine 2021;384(15):1402–11.

17. Power MJ, R. K, Mc Guffin DC, Lam D, Beck AT. The Dysfunctional Attitude Scale (DAS): A comparison of Forms A and B and proposal for a new subscale version. Journal of Research in Personality. 1994;28:263–76.

18. Scheier MF, Carver CS, Bridges MW. Distinguishing optimism from neuroticism (and trait anxiety, self-mastery and self-esteem): A re-evaluation of the Life Orientation Test. Journal of Personality and Social Psychology. 1994;67:1063–78.

19. Weiss B, Erritzoe DE, Giribaldi B, Nutt DJ, Carhart-Harris RL. A critical evaluation of QIDS-SR-16 using data from a trial of psilocybin therapy versus escitalopram treatment for depression. Journal of Psychopharmacology (Oxford, England). 2023;37(7):717–32.

20. Diener E, Wirtz D, Tov W, Kim-Prieto C, Choi D, Oishi S, et al. New measures of well-being: Flourishing and positive and negative feelings. Social Indicators Research. 2009;39:247–66.

21. Pedrelli P, Feldman GC, Vorono S, Fava M, Peterson T. Dysfunctional attitudes and perceived stress predict depressive symptoms severity following antidepressant treatment in patients with chronic depression. Psychiatry Research. 2008;161(3):302–8.

22. Grob CS, McKenna DJ, Callaway JC, Brito GS, Neves ES, Oberlaender G, et al. Human psychopharmacology of hoasca, a plant hallucinogen used in riutal context in Brazil. The Journal of Nervous and Mental Disease 1996;184(2):86–4.

23. Goodwin GM, Price J, De Bodinat C, Laredo J. Emotional blunting with antidepressant treatments: a survey among depressed patients. Journal of Affective Disorders. 2017;221:31–5.

24. McCabe C, Mishor Z, Cowen PJ, Harmer CJ. Diminished neural processing of aversive and rewarding stimuli during selective serotonin reuptake inhibitor treatment Biological Psychiatry. 2010;67(5) 439–45.

25. Gloster AT, Gerlach AL, Hamm A, Höfler M, Alpers GW, Kircher T, et al. 5HTT is associated with the phenotype psychological flexibility: results from a randomized clinical trial. European Archives of Psychiatry and Clinical Neuroscience. 2015;265(5) 399–406.

26. Frokjaer VG, Vinberg M, Erritzoe DE, Baaré W, Holst KK, Mortensen EL, et al. Familial risk for mood disorder and the personality risk factor, neuroticism, interact in their association with frontolimbic serotonin 2A receptor binding. Neuropsychopharmacology: Official Publication of the American College of Neuropsychpharmacology. 2010;35(5):1129–37.

27. Carhart-Harris RL, Friston KL. REBUS and the Anarchic Brain: Toward a Unified Model of the Brain Action of Psychedelics. Pharmacology Reviews. 2019;71(3):316–44.

28. Alamia A, Timmerman C, Nutt DJ, VanRullen R, Carhart-Harris RL. DMT alters cortical travelling waves. Elife. 2020;9.

29. Bogenschutz MP, Forcehimes AA, Pommy JA, Wilcox CE, Barbosa PCR, Strassman RJ. Psilocybin-assisted treatment for alcohol dependence: a proof-of-concept study. Journal of Psychopharmacology (Oxford, England). 2015;29(3):289–99.

30. Moreno FA, Weigand CB, Taitano EK, Delgado PL. Safety, tolerability, and efficacy of psilocybin in 9 patients with obsessive-compulsive disorder. The Journal of Clinical Psychiatry. 2006;67(11):1735–40.

